# *APOE4* and Chronic Health Risk Factors are Associated with Sex-Specific Preclinical Alzheimer’s Disease Neuroimaging Biomarkers

**DOI:** 10.1101/2024.11.21.24317732

**Authors:** Genna M. Losinski, Mickeal N. Key, Eric D. Vidoni, Jonathan Clutton, Jill K. Morris, Jeffrey M. Burns, Amber Watts

## Abstract

**Introduction:** Two thirds of Alzheimer’s disease (AD) patients are female. Genetic and chronic health risk factors for AD affect females more negatively compared to males.

**Objective:** This exploratory multimodal neuroimaging study aimed to examine sex differences in cognitively unimpaired older adults on: (1) amyloid-β via 18F-AV-45 Florbetapir PET imaging, (2) neurodegeneration via T1 weighted MRI volumetrics, (3) cerebral blood flow via ASL-MRI. We identified AD risk factors including genetic (*APOE* genotype status) and health markers (fasting glucose, mean arterial pressure, waist-to-hip ratio, and android and gynoid body fat) associated with neuroimaging outcomes for which we observed sex differences.

**Methods:** Participants were sedentary, amyloid-β positive older adults (N = 112, ages 65-87 years) without evidence of cognitive impairment (CDR = 0).

**Results:** Multivariate analysis of covariance models adjusted for intracranial volume, age, and years of education demonstrated lower volume (*F* (7, 102) = 2.67, p = 0.014) and higher blood flow *F* (6, 102) = 4.25, p =<0.001) among females compared to males in regions of interest connected to AD pathology and the estrogen receptor network. We did not observe sex differences in amyloid-β levels. Higher than optimal waist to hip ratio was most strongly associated with lower volume, while higher android fat percentage and *APOE* ε4 carrier status were most strongly associated with higher blood flow among female participants. Discussion: Findings suggest genetic and chronic health risk factors are associated with sex-specific AD neuroimaging biomarkers. Underlying sex-specific biological pathways may explain these findings. Our results highlight the importance of considering sex differences in neuroimaging studies and when developing effective interventions for AD prevention and risk reduction.

## 1.0 INTRODUCTION

The female sex has long been established as a major risk factor for late-onset Alzheimer’s disease (AD) [1]. Two thirds of patients with AD are women [2]. Sex differences reported in the incidence and prevalence of AD have historically been attributed to women’s longer life expectancy relative to men [3,4]. Further, research examining AD risk factors has historically adjusted for sex as a covariate in analyses but has given limited attention to describing or explaining sex and gender differences. Recent research suggests sex and gender differences in AD risk factors may better explain disparities in prevalence and has identified approximately 30 AD risk factors that affect men and women differently, with the female sex often more severely affected [5–7].

The prevalence rates of several AD risk factors (e.g., cerebrovascular events, depression, sleep disorders) are higher among women, especially after the age of 60 and during the postmenopausal period [8–13]. One hypothesis proposed to explain this sex disparity suggests estrogen deprivation, as is observed in the menopause transition, may function as a physiological trigger of increased risk for AD given estrogen’s (specifically 17b-estradiol) neuroprotective properties for the female brain [7,14–17]. The estrogen hypothesis emphasizes the numerous fundamental functions of estrogen highlighting an extensive network of estrogen receptors throughout the female brain, predominantly in the hypothalamus, hippocampus, and amygdala [18,19], areas of the brain known to be associated with early AD pathophysiological changes. The most robust evidence for sex-differences in AD risk are found in genetic risk factors (i.e., Apolipoprotein E (*APOE*) genotype) and chronic health factors (e.g., cardiometabolic disease) [5].

The most studied and strongest genetic risk factor for late onset AD is *APOE* genotype status [20,21]. *APOE* gene expression has been shown to be more detrimental for female carriers of the *APOE4* allele. Females that carry the *APOE4* allele have 1.5 times higher risk for AD [1,22,23] and more amyloid-β plaques and neurofibrillary tangles compared to male *APOE4* carriers [20]. The literature has suggested an age-specific effect in females wherein female *APOE4* carriers may have an early susceptibility to AD compared to males [24]. Among cognitively intact older adult *APOE4* carriers, females have greater decreases in hippocampal connectivity, and increased whole brain hypometabolism and atrophy compared to age matched male *APOE4* carriers [25,26]. The mechanisms underlying the interaction between sex and *APOE* genotype remain unclear.

Cardiometabolic diseases (e.g., type 2 diabetes, metabolic syndrome) and their risk factors (e.g. hypertension, hyperlipidemia, non-optimal fasting glucose, obesity) associated with sedentary behavior are well-known risk factors for AD development and progression for both men and women [27–30]. The development, symptoms, and treatment of cardiometabolic diseases differ by sex [6,31–35]. For example, microvascular disease contributes the most to cardiovascular disease in women compared to obstructive coronary artery disease in men [36]. Additionally, men tend to develop cardiometabolic disease at an earlier age than women given that the menopause transition and loss of neuroprotective estrogen may accelerate the development of cardiometabolic diseases in postmenopausal women [37–39]. Estrogen has demonstrated a protective effect on the brain through its vasodilatory effects, enhancing the production of sensitivity to vasodilatory factors [40,41]. Historically, the literature has suggested a greater impact of vascular disease for men’s AD risk versus women, yet given the growing body of literature on sex differences and cardiometabolic disease, it has been suggested that the diagnosis of hypertension, high cholesterol, and diabetes may put women at a higher risk for AD [6]. However, more research is needed.

Emerging lines of evidence suggest one out of every three AD cases can be linked to modifiable risk factors [42]. Thus, early detection and prevention efforts have dominated the field in addressing the AD epidemic. Given that many of these identified cardiometabolic disease risk factors are modifiable, it is important to fully understand sex differences, including how they contribute to AD-specific pathophysiological changes (amyloid-β accumulation, neurofibrillary tangles, and neuronal and synaptic loss) when developing effective AD risk reduction interventions. Through various imaging techniques, early detection of amyloid, tau, and neurodegeneration (A/T/N) biomarkers that begin in the brain before detectable clinical symptoms, can identify cognitively unimpaired individuals that may be at high risk for developing AD, and therefore benefit the most from interventions to prevent or delay the onset of AD.

When considering the evidence of amyloid-β accumulation beginning as early as 15-20 years before clinically observable cognitive decline and age being the strongest predictor for AD, this suggests amyloid-β accumulation may begin in or before midlife, which notably coincides with the menopause transition for women and the inherent loss of the neuroprotective 17b-estradiol [16,43,44]. New evidence indicates the presence of sex differences among AD-related biomarkers start in midlife such that menopausal women exhibit more amyloid-β accumulation and decreased gray and white matter volumetrics among AD-specific regions of the brain compared to age-matched men [45–47]. However, the research examining whether these early AD biomarker sex differences persist with aging among cognitively unimpaired older adults with consideration to postmenopausal women, remains mixed. Further, in-vivo multi-modal imaging studies have shown 17b-estradiol densities among estrogen-regulated networks vary over the menopause transition, suggesting women’s menopausal status is important to interpretation of neuroimaging results [48].

Studies that have examined sex differences in amyloid-β accumulation among cognitively unimpaired older adults have shown conflicting results including no sex differences [49,50], older adult males exhibiting more amyloid-β accumulation [51], and older adult females exhibiting more amyloid-β accumulation [52]. Sex differences among volumetric studies indicate cognitively unimpaired older adult males exhibit more age-related atrophy in the temporal regions of the brain compared to older adult women, including smaller hippocampus volume indicative of possible downstream AD neurodegeneration [49,53,54]. However, review of longitudinal studies examining brain atrophy, amyloid-β accumulation, and cognitive decline have demonstrated sex differences in the progression of AD such that women may be protected relative to men during the prodromal AD phases, but exhibit faster progression rates of decline compared to men [5,55].

In addition to the typical imaging techniques and biomarkers used to detect early pathophysiological processes of AD following the A/T/N classification system, decreased cerebral blood flow has been discussed as a novel AD biomarker given the hypothesis of vascular abnormality of AD in conjunction with the ability of ASL-MRI to detect tissues and cell damage that precede neurodegeneration [56–59]. Further, cerebral blood flow is an important index of aging and it is well known that velocity decreases as a function of aging [60–62].

Research has demonstrated sex differences in cerebral blood flow. An increased rate of cerebral blood flow in young adult women compared to age-matched men has been widely reported and observed using a variety of types of techniques (e.g. single photon emission computed tomography (SPECT), PET, Xenon-enhanced computed tomography, and ASL-MRI) [63–67]. Few studies have examined sex differences in cerebral blood flow among older adults and those few report conflicting results [68,69]. One unique cross-sectional study looked at the rate of changes in cerebral blood flow across the lifespan in women and men [70]. Results indicated sex-specific trajectories with aging such that women had a greater rate of decline of cerebral blood flow over the lifespan compared to men and that this disparity in the declining rate was most pronounced during ages 61-70 years old [70]. For women, cerebral blood flow has been found to differ dependent on menopause stage, such that postmenopausal women demonstrate higher cerebral blood flow compared to other menopause stages (i.e., pre- and peri-menopausal) in certain brain regions [71].

Taken together, this research suggests contradictory results regarding sex differences in amyloid-β accumulation, AD related brain volumes, and cerebral blood flow in cognitively unimpaired older adults. These discrepancies suggest that there may be confounding variables that have yet to be fully explored, such as specific risk factors that may contribute to the findings and the timing of data collection relative to aging and menopause processes. For example, recent studies suggest that brain volume and connectivity change across the menstrual cycle [72,73] and estrogen receptor density changes in estrogen regulated networks throughout the menopause transition [48], though these cycles are rarely considered in neuroimaging research. The underlying mechanistic explanations for these observed sex differences remain understudied. The role of sex hormones has been postulated as a potential contributing factor to these results [41] and examining the associations with sex specific risk factors may help with clarification. For example, Rahman et al. [44] uniquely examined the relationship between sex-specific risks and AD biomarkers in a cohort of middle-aged adults. The study demonstrated higher amyloid-β deposition and lower MRI gray and white matter volumes in middle-aged women compared to age-matched men. Notably, hormonal risk factors, including menopause stage and hormone replacement therapy use, predicted these identified sex differences among the biomarkers when compared to other clinical, chronic health, and lifestyle AD risk factors [47].

The present study sought to investigate sex differences in the association of brain volume and cerebral blood flow with chronic health risk factors in a cohort of sedentary, amyloid-β positive, older adults. Our multimodal imaging study examined in vivo sex differences in a cohort of high-risk, cognitively unimpaired older adults on the following AD biomarkers: (1) amyloid-β on 18F-AV-45 Florbetapir PET imaging, (2) neurodegeneration via TI weighted MRI volumetrics, and (3) cerebral blood flow via ASL-MRI in AD-specific brain regions. Moreover, we aimed to identify which of the sex-specific AD risk factors (genetic, cardiometabolic health factors) was most strongly associated with the sex-driven AD brain biomarker differences. Based on the estrogen hypothesis, we hypothesized that cognitively unimpaired older adult women would exhibit greater amyloid-β burden, reduced volume, and reduced cerebral blood flow in preclinical AD-specific brain regions (e.g. amygdala, hypothalamus, and parahippocampal gyrus) compared to men and that genetic (*APOE* genotype) and health risk factors (cardiometabolic disease) would predict these differences. Findings contribute to understanding the optimal timing of AD prevention trials while gaining insight regarding AD risk in the postmenopausal stage for women.

## 2.0 MATERIALS AND METHODS

### 2.1 PARTICIPANTS

The present study is a secondary analysis utilizing the baseline data from the Alzheimer’s Prevention through Exercise (APEX, NCT02000583) study, a 52-week exercise intervention conducted at the University of Kansas Alzheimer’s Disease Research Center examining change in AD-related neuroimaging biomarkers. Baseline medical history, neuropsychological testing, and MRI and amyloid-β imaging with PET (18F-AV-45) were assessed in all participants during baseline evaluations. The APEX study recruited a convenience sample of 121 cognitively unimpaired, sedentary older adult participants. Inclusion criteria for APEX required age of 65 years and older, a Clinical Dementia Rating (CDR) Scale of 0, a Mini Mental State Examination (MMSE) score of 27 or greater, normal cognitive test performance on the Uniform Data Set neuropsychiatric battery [74,75] for age and years of education (<1.5 SD below the mean), stable 30-day medication regimen, sedentary or underactive as defined by the Telephone Assessment of Physical Activity [76], and amyloid-β positive measured by PET (18F-AV-45). Persons with insulin-dependence, significant hearing or vision problems, clinically evident stroke, cancer in the previous 5 years, or recent history (<2 years) of major cardiorespiratory, musculoskeletal, or neuropsychiatric impairment, were excluded.

### 2.2 GENETIC RISK FACTORS

*APOE* genotype status was determined from frozen whole blood samples (acid citrate dextrose anticoagulant) using an allelic discrimination assay to identify single nucleotide polymorphisms (ThermoFisher). We distinguished *APOE4*, *APOE3*, and *APOE2* alleles using Taqman probes to APOE-defining polymorphisms: rs429358 (C_3084793_20) and rs7412 (C_904973_10). This was included in the model as the number of ε4 alleles (0, 1, 2).

### 2.3 CHRONIC HEALTH RISK FACTORS

The presence or absence of the following cardiovascular risk factors were included as predictive measures in the present analysis: 1) Optimal fasting glucose: A fasting glucose measurement was collected twice during the baseline visit and an average of these measurements was calculated. A fasting glucose level of above 100 mg/dL was considered non-optimal [77]. 2) High mean arterial pressure: Mean arterial pressure was collected at the baseline visit and pressures that read above 100 was considered high [78]. 3) Higher than optimal waist to hip ratio: Waist to hip ratio was calculated at the baseline visit and dichotomized as yes/no depending on whether it was above the sex-specific cutoff of greater than or equal to 0.85 in women and greater than or equal to 0.90 in men [79]. A Dual-energy X-ray absorptiometry (DEXA) scan was completed during the baseline visit to measure total body, android (trunk and abdomen adipose tissue), and gynoid (hip and thigh adipose tissue) fat percentages. 4) Higher than optimal total body fat percentage: Total body fat percentage was calculated by dividing fat mass (g) by the sum of lean mass (g), bone mineral content (g), and fat mass (g). Total body fat percentage greater than 38% in women and greater than 25% in men was considered nonoptimal [80]. 5) Total android fat percentage: Android fat percentages were calculated by dividing android fat (g) by total fat mass (g). 6) Total gynoid fat percentage: Gynoid fat percentages were calculated by dividing gynoid fat (g) by total fat mass (g).

### 2.4 AD BRAIN BIOMARKER IMAGING

For the proposed analysis, sex differences were examined among three neuroimaging techniques (18F-AV-45 PET imaging, T1 weighted MRI, and ASL-MRI, described in detail below).

#### 2.4.1 AMYLOID-β PET ACQUISITION, PROCESSING, AND ANALYSIS

All participants received Florbetapir PET scans obtained approximately 50 minutes after administration of intravenous florbetapir 18F-AV45 (370 MBq) acquired with the GE Discovery ST-16 PET/CT scanner. Two PET brain frames of five minutes in duration were acquired continuously, summed, and attenuation corrected. The mean PET signal across the brain was divided by the signal from whole cerebellum ROI to produce a Standardized Uptake Ratio (SUVR) image. Three experienced raters interpreted the images to determine amyloid status as “elevated” or “non-elevated” as previously described [81–84] . Raters first reviewed raw PET images visually then examined the cerebellum normalized SUVRs in 6 cortical regions (anterior cingulate, posterior cingulate, precuneus, inferior medial frontal, lateral temporal, and superior parietal cortex) and projection maps comparing SUVRs to an atlas of amyloid negative scans [83]. Participants for the primary exercise trial were eligible if they had an “elevated” scan or were in the “subthreshold” range which was defined as a mean cortical SUVR for the 6 ROIs > 1.0, which represented the upper half of non-elevated scans (mean cortical SUVR for non-elevated scans [n = 166] 0.99 [0.06 SD]) [81]. Therefore, participants included in the present secondary analysis were determined to have “sub-threshold” and “elevated” amyloid status as determined from the primary exercise trial. To remain consistent with universal quantitative standards, SUVR values were converted to Centiloid values (Centiloids = 196.9 × SUVR_wholecerebrefFBP − 196.03) [85,86]. The Centiloid values for the sample met the ADNI standard amyloid-β positivity threshold (25.3 Centiloids/composite reference region). Therefore, participants included in the present secondary analysis were determined “amyloid-β positive.”

#### 2.4.2 MRI ACQUISITION, PROCESSING, AND ANALYSIS

All participants received Siemens Skyra 3T T1-weighted MRI of the brain (Tesla Skyra scanner; MP-RAGE 1 × 1 × 1.2 mm voxels, TR = 2300 ms, TE = 2.98 ms, TI = 900 ms, FOV 256 × 256×mm, 9° flip angle; Pulsed ASL single-shot EPI 3.8 × 3.8 × 4.0 mm,TR = 3400 ms, TE = 13 ms, TI = 700 ms, FOV 240 × 240×mm, 90° flip angle; ASL single-shot EPI 3.8×3.8×4.0mm,TR = 3400ms, TE = 13ms, TI = 700ms, FOV 240×240mm, 90° flip angle). To facilitate assessment across imaging modalities, participants’ sequences were co-registered to the anatomical native space using SPM12 (https://www.fil.ion.ucl.ac.uk/spm/software/spm12). The anatomical image was segmented using CAT12 and the Neuromorphometic atlas (https://neuro-jena.github.io) producing both regional volumes and region of interest (ROI) masks. These individualized ROI were used to extract mean values from the ASL-MRI and 18F-AV-45 PET SUVR images. ASL-MRI data were processed using the ASLTbx for SPM12 before mean blood flow values were extracted from each ROI.

#### 2.4.3 BRAIN REGIONS OF INTEREST

Bilateral ROIs examined via 18F-AV-45 Florbetapir PET imaging included the anterior cingulate gyrus, lateral temporal lobe, precuneus, and superior parietal lobe. The anterior cingulate gyrus was primarily examined based on the results of the Rahman et al. 2020 study that found amyloid-β deposition sex differences in this region among middle-aged adults. The additional neocortical areas (lateral temporal lobe, precuneus, and superior parietal lobe) in addition to a global measurement of amyloid-β deposition were included because of their identified association with preclinical AD according to the amyloid cascade hypothesis of AD [87,88].

Bilateral ROIs examined via TI weighted MRI volumetrics included the hippocampus, amygdala, parahippocampal gyrus, entorhinal cortex, insula, and caudate. The hippocampus, amygdala, parahippocampal gyrus, ínsula, and caduate were primarily examined based on the findings of the Rahman et al. 2020 study that demonstrated sex differences among these 5 regions in a cohort of middle-aged adults. The entorhinal cortex was also included as an exploratory ROI because it has been identified among the literature as one of the earliest brain regions found to be associated with Preclinical Alzheimer’s disease via structural MRI, along with the hippocampus, amygdala, and parahippocampal gyrus [89]. The hippocampus and the amygdala are also brain regions included in the estrogen receptor network and therefore considered exploratory ROI for the present study examining sex differences [90].

Bilateral ROIs examined via MRI-ASL included the hippocampus, amygdala, parahippocampal gyrus, temporal lobe, precuneus, anterior cingulate cortex, and superior parietal lobe. The precuneus, anterior cingulate cortex, and superior parietal lobe were primarily examined based on research that has found reduced cerebral blood flow in these three regions in patients with AD [91–94]. The hippocampus, amygdala, parahippocampal gyrus, and temporal lobes were included as exploratory regions due to their association with preclinical AD [89,90].

Bilateral ROIs examined via 18F-AV-45 Florbetapir PET imaging included the anterior cingulate gyrus, lateral temporal lobe, precuneus, and superior parietal lobe. The anterior cingulate gyrus was primarily examined based on the results of the Rahman et al. 2020 study that found amyloid-β deposition sex differences in this region among middle-aged adults. The additional neocortical areas (lateral temporal lobe, precuneus, and superior parietal lobe) in addition to a global measurement of amyloid-β deposition were included because of their identified association with preclinical AD according to the amyloid cascade hypothesis of AD [87,88].

### 2.4 STATISTICAL ANALYSES

#### 2.4.1 AIM 1: AD BRAIN BIOMARKER DIFFERENCES BY SEX

To investigate regional differences between men and women in T1 weighted volumes (MRI), amyloid-β deposition (18F-AV-45 PET), and cerebral blood flow (ASL-MRI), we applied the following three Multivariate Analysis of Covariance (MANCOVA) models to examine whether regions differed by sex while controlling for age, education, and modality specific confounders (TIV = total intracranial volume, number of *APOE4* alleles).

MANCOVA 1: Amyloid-β deposition (18F-AV-45 PET) of 5 ROIs ∼ Sex + Age + Education + *APOE4* status

MANCOVA 2: T1 weighted volumes (MRI) of 6 ROIs ∼ Sex + Age + Education + TIV MANCOVA 3: Cerebral blood flow (ASL-MRI) of 7 ROIs ∼ Sex + Age + Education

Univariate ANCOVAS were used to further examine significant group differences followed by Tukey’s HSD Test for multiple comparisons.

#### 2.4.2 AIM 2: ASSOCIATIONS BETWEEN SEX-DRIVEN AD BRAIN BIOMARKER DIFFERENCES AND AD RISK FACTORS (GENETIC, CHRONIC HEALTH CONDITIONS)

To identify which of the risk factor variables were most strongly associated with sex differences in neuroimaging biomarkers identified in Aim 1, we used Least Absolute Shrinkage and Selection Operator (LASSO) regressions, controlling for age and education. The LASSO regression method was selected due to the high levels of multicollinearity among the predictors (Table 1), the combination of different types of predictors (i.e., continuous versus discrete), and to exclude extraneous variables and obtain a subset of predictors that are, empirically, the strongest associations with the sex-related biomarker differences in our models [95]. Given the sample size and number of predictors, LASSO regressions within a k 2-fold cross validation using a leave one out predictive modeling framework were applied to the data to choose final models with the lowest minimum cross-validated error and identify the most essential predictors.

**Table 1.** Correlation Matrix: Pearson correlation coefficients (for continuous variables) / Point biserial coefficient (for one continuous variable and one dummy variable) / Phi coefficient (for two dummy variables)

| Variable | 1 | 2 | 3 | 4 | 5 | 6 | 7 | 8 | 9 | 10 |
| --- | --- | --- | --- | --- | --- | --- | --- | --- | --- | --- |
| 1 Age (years) | 1 |  |  |  |  |  |  |  |  |  |
| 2 Education (years) | 0.02 | 1 |  |  |  |  |  |  |  |  |
| 3 Sex (male/female) | -0.03 | -0.14 | 1 |  |  |  |  |  |  |  |
| 4 *APOE4* status<br>(0,1 alleles) | 0.06 | -0.02 | 0.01 | 1 |  |  |  |  |  |  |
| 5 Fasting plasma glucose (yes/no) | -0.05 | -0.13 | 0.21 | 0.01 | 1 |  |  |  |  |  |
| 6 Mean arterial pressure (yes/no) | -0.12 | -0.09 | 0.01 | 0.07 | **0.44** | 1 |  |  |  |  |
| 7 Waist-to-hip ratio (yes/no) | -0.04 | 0.08 | **1.99** | 0.01 | **0.46** | 0.01 | 1 |  |  |  |
| 8 Total body fat (yes/no) | 0.07 | **0.22** | **2.36** | 0.01 | 0.02 | 0.01 | **0.82** | 1 |  |  |
| 9 Android fat, % | -0.04 | -0.01 | 0.15 | -0.08 | 0.02 | 0.15 | -0.05 | -0.15 | 1 |  |
| 10 Gynoid fat, % | 0.02 | 0.09 | -0.14 | 0.12 | 0.05 | -0.01 | 0.05 | 0.24 | **-0.74** | 1 |

Variables that were included as potential predictors include: *APOE4* status (0, 1, 2 alleles), non-optimal fasting glucose (yes/no), elevated mean arterial pressure (yes/no), higher than optimal waist to hip ratio (yes/no), higher than optimal body fat percentage (yes/no), total android fat (percentage), and total gynoid fat (percentage). Covariates in this analysis included: age (years), and education (years). To identify the risk factors specific to sex, interaction terms between the sex variable and the risk factor variables were also included.

## 3.0 RESULTS

### 3.1 PARTICIPANTS

Data from 121 participants were available for analysis. We excluded 9 participants (6 with incomplete genetic testing data and 3 with incomplete MRI data). The remaining 112 participants (ages 65-87 years) were examined, including 74 women and 38 men who were college educated on average and 95.5% of which identified as non-Hispanic, White. Results from the Community Healthy Activities Model Program for Seniors (CHAMPS) self-report questionnaire suggest the sample did not engage in a substantial amount of physical activity and were primarily sedentary. Participants’ clinical, demographic, and anthropometric characteristics are given in Table 2. Males and females did not differ by age, *APOE4* carrier status, fasting plasma glucose, mean arterial pressure, or physical activity engagement. The female group was less educated (*p* = 0.049) and included a higher percentage of participants who identified as African American (*p* < 0.001), had a lower waist to hip ratio (*p* <0.001), lower total body fat percentage (*p* < 0.001), higher android fat percentage (*p* <0.001), and lower gynoid fat percentage (*p* <0.001) than the male group.

**Table 2.**
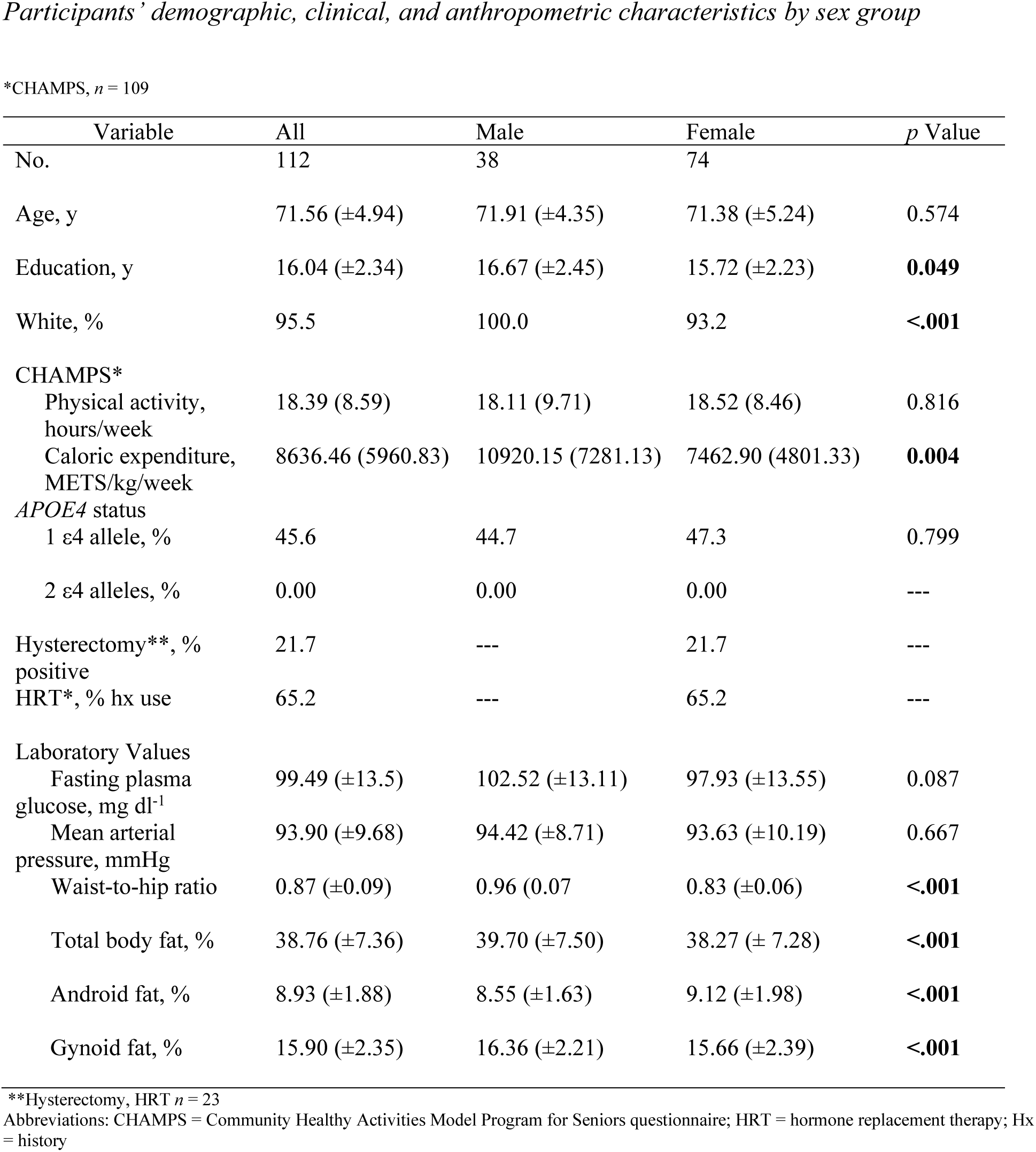
Participants’ demographic, clinical, and anthropometric characteristics by sex group

### 3.2 AIM 1:AD BRAIN BIOMARKER DIFFERENCES BY SEX

Brain ROIs showing biomarker differences between sex groups were first identified among amyloid-β deposition (18F-AV-45 PET), T1 weighted volumetrics (MRI), and cerebral blood flow (ASL-MRI) among the whole sample.

#### 3.2.1 PET AMYLOID-β BURDEN

A Multivariate Analysis of Covariance (MANCOVA) of the relationship between sex and amyloid-β burden (Centiloid) in four hypothesized brain ROIs (anterior cingulate gyrus, lateral temporal lobe, precuneus, superior parietal lobule) and global amyloid-β burden adjusting for age, education, and *APOE4* status revealed a non-significant trend toward group differences between men and women (*F* (5, 103) = 2.09, p = 0.071), and post hoc individual ANCOVAs revealed no significant group differences for the these ROIs (Table 3).

**Table 3.** Sex group differences by PET imaging modality (centiolid)

| Region | All (n=112) | Male (n=38) | Female (n=74) | *p* Value |
| --- | --- | --- | --- | --- |
| Anterior Cingulate Gyrus | 60.99 (±41.05) | 60.04 (±43.75) | 61.48 (±39.89) | 0.853 |
| Lateral Temporal Lobe | 45.84 (±33.99) | 47.56 (±32.66) | 44.96 (±34.92) | 0.853 |
| Precuneus | 58.02 (±44.75) | 61.13 (±48.58) | 56.43 (±42.90) | 0.853 |
| Superior Parietal Lobule | 26.91 (±34.86) | 22.84 (±32.96) | 28.99 (±35.83) | 0.667 |
| Global Amyloid Burden | 1240.14 (±206.18) | 1245.85 (±217.95) | 1237.22 (±201.33) | 0.944 |

Given the unequal sex sample sizes in this analysis (males = 38, females = 74), the Box’s Test for Equivalence of Covariance Matrices (Box’s M) was conducted and interpreted using p < .001 as a criterion [96,97]. The Box’s M test was not significant (*p* = 0.120), suggesting the assumption of homogeneity of covariance matrices was met.

#### 3.2.2 MRI VOLUMES

After adjusting for intracranial volume, age, and years of education, a MANCOVA yielded a significant group difference between male and female volumes in six hypothesized ROIs (*F* (6, 102) = 4.25, p <0.001) (Table 4). The Box’s M test was not significant (*p* = 0.115), suggesting the assumption of homogeneity of covariance matrices was met. Post hoc univariate ANCOVAs, corrected with the Benjamini-Hochberg critical value for a false discovery rate of 5%, among the six hypothesized ROIs revealed there was a statistically significant difference in volumes between males and females in the amygdala (*F* (1, 107) = 4.78, *p* < 0.048) and entorhinal cortex (*F* (1, 107) = 15.89, *p* <=0.002). Tukey’s HSD Test for multiple comparisons found that females exhibited lower volume in the amygdala (*p* < 0.048, 95% C.I. = -0.01, 0.09), and entorhinal cortex (*p* < 0.002, 95% C.I. = 0.07, 0.33) compared to males (Figure 1).

**Table 4.** Sex group differences by MRI imaging modality

| Modality | All (n=112) | Male (n=38) | Female (n=74) | p Value |
| --- | --- | --- | --- | --- |
| MRI-Volume*<br>Region, mL |  |  |  |  |
| Hippocampus | 6.40 (±0.65) | 6.74 (±0.70) | 6.22 (±0.55) | 0.884 |
| Amygdala | 1.74 (±0.18) | 1.86 (±0.18) | 1.68 (±0.14) | **0.048** |
| Parahippocampal gyrus | 6.75 (±0.66) | 7.22 (±0.71) | 6.51 (±0.49) | 0.536 |
| Entorhinal cortex | 4.28 (±0.48) | 4.66 (±0.44) | 4.08 (±0.37) | **0.002** |
| Insula | 11.73 (±1.13) | 12.41(±1.24) | 11.38(±0.90) | 0.603 |
| Caudate | 5.67 (±0.66) | 5.93 (±0.73) | 5.53 (±0.57) | 0.205 |
| MRI-ASL<br>Region, mL/100g/min |  |  |  |  |
| Amygdala | 28.57 (±) 8.25 | 25.13 (±) 7.76 | 30.33 (±) 7.94 | **0.014** |
| Temporal Lobe | 31.18 (±) 6.57 | 28.20 (±) 5.90 | 32.71 (±) 6.30 | **0.005** |
| Anterior Cingulate Cortex | 23.45 (±) 6.75 | 20.74 (±) 6.04 | 24.84 (±) 6.31 | **0.014** |
| Precuneus | 28.73 (±) 8.08 | 24.73 (±) 7.79 | 30.78 (±) 7.38 | **0.005** |
| Superior Parietal Lobe | 21.70 (±) 6.28 | 19.02 (±) 6.77 | 23.07 (±) 5.64 | **0.005** |
| Parahippocampal Gyrus | 31.60 (±) 8.87 | 28.22 (±) 9.04 | 33.34 (±) 8.26 | **0.015** |
| Hippocampus | 31.76 (±) 7.94 | 28.23 (±) 6.80 | 33.57 (±) 7.37 | **0.005** |

\*adjusted for total intracranial volume

**Figure 1.**
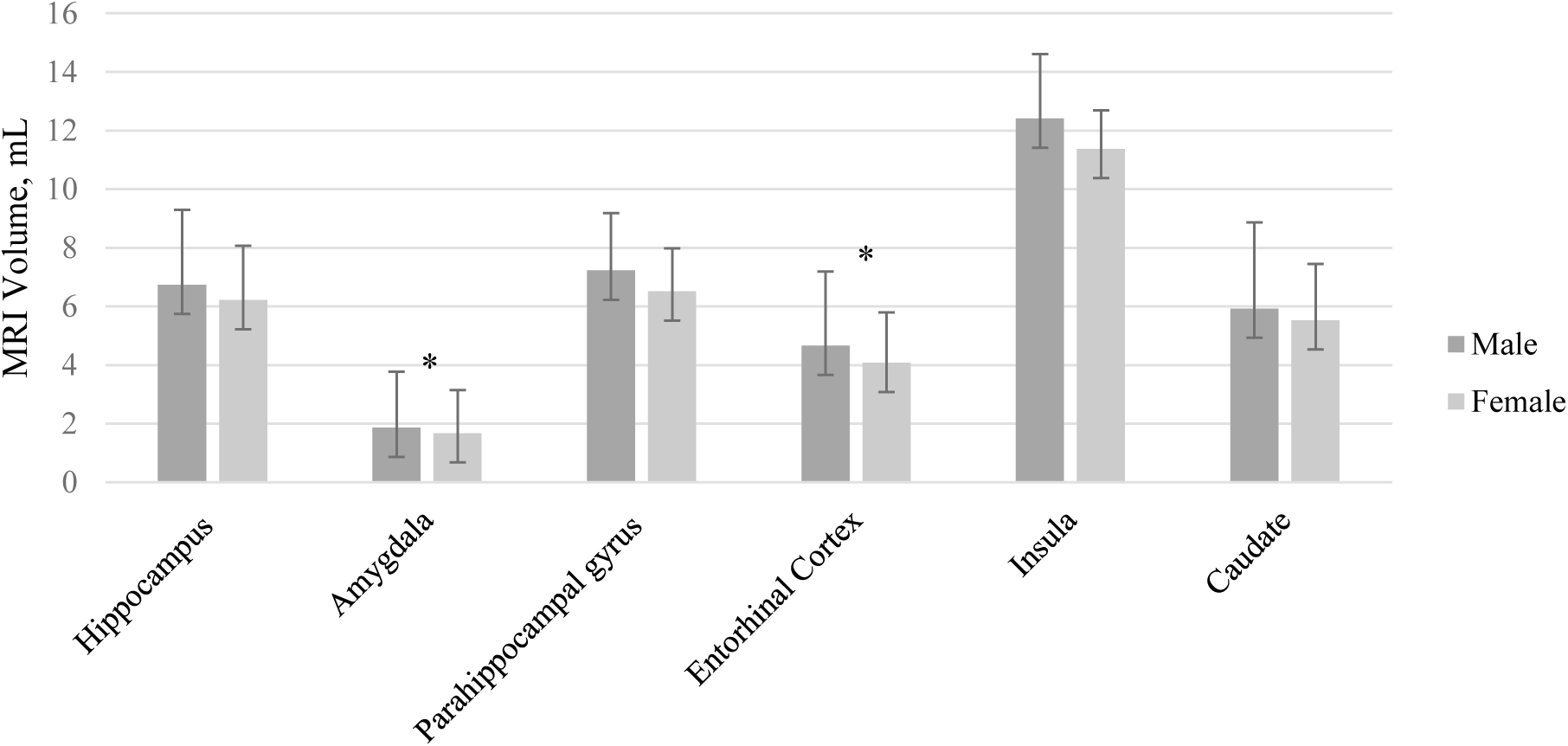
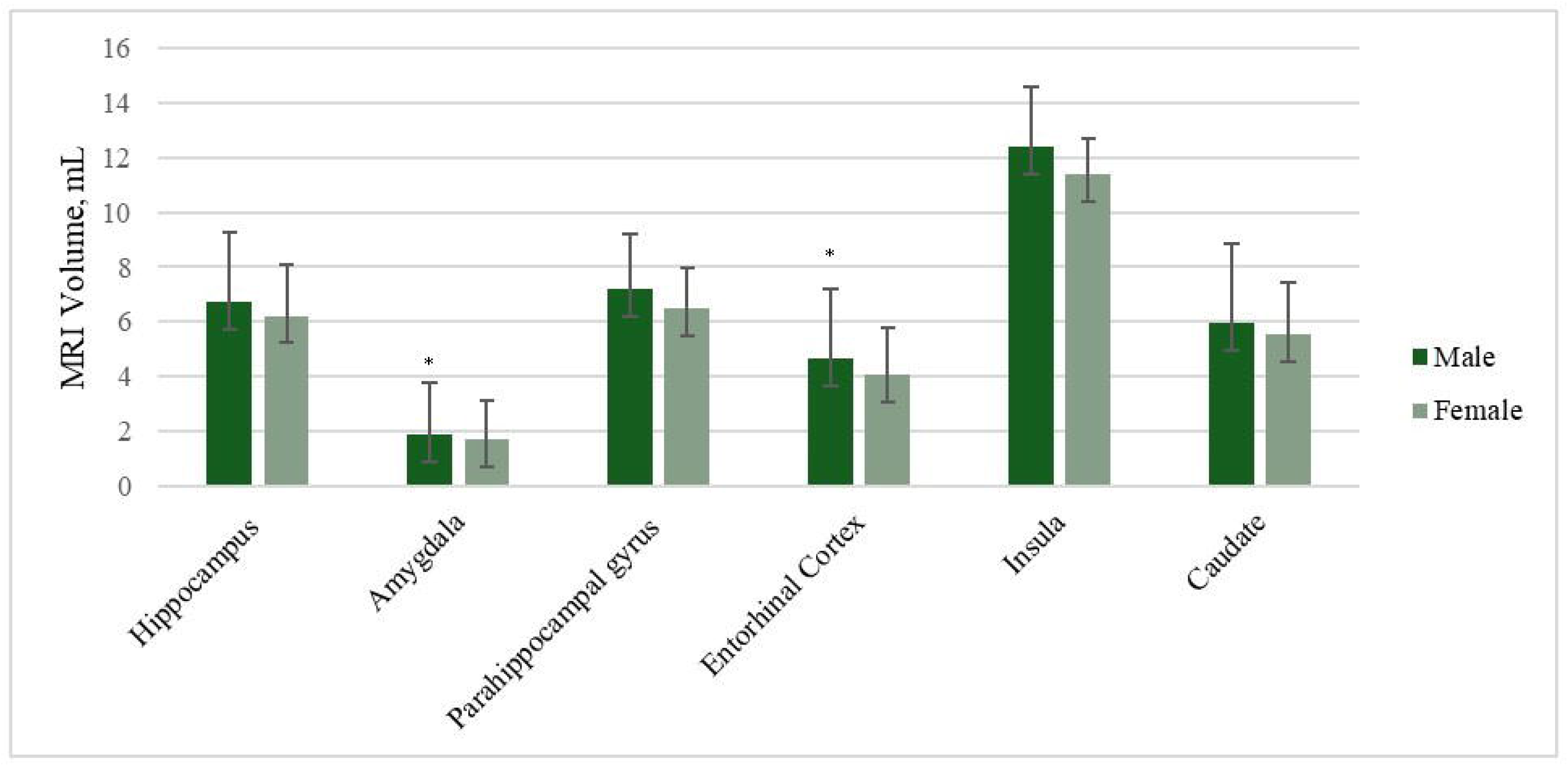
MRI Volume Sex Differences by Region of Interest accounting for differences in total volume, age, and education. Error bars represent 95% confidence intervals.

#### 3.2.3 ASL-MRI CEREBRAL BLOOD FLOW

A MANCOVA of the relationship between sex and cerebral blood flow in hypothesized brain ROIs after adjusting for age and years of education was statistically significant, and the female group showed higher blood flow compared to the male group in several brain regions (*F* (7, 102) = 2.67, p = 0.014) (Table 4). Given the unequal sex sample sizes in this analysis (males = 38, females = 74), the Box’s Test for Equivalence of Covariance Matrices (Box’s M) was conducted and interpreted using p < .001 as a criterion [96,97]. The Box’s M test was not significant (*p* = 0.002), suggesting the assumption of homogeneity of covariance matrices was met. Post hoc univariate ANCOVAs, corrected with the Benjamini-Hochberg critical value for a false discovery rate of 5%, among the seven hypothesized ROIs revealed there was a statistically significant difference in cerebral blood flow between males and females in the amygdala (*F* (1, 108) = 8.70, *p* = 0.014), temporal lobe (*F* (1, 108) = 12.46, *p =* 0.005), anterior cingulate cortex (*F* (1, 108) = 8.65), *p* = 0.014), precuneus (*F* (1, 108) = 17.29, *p* = 0.005), superior parietal lobe (*F* (1, 108) = 11.93, *p* = 0.005), parahippocampal gyrus (*F* (1, 108) = 8.26, *p* = 0.015), and hippocampus (*F* (1, 108) = 11.66, p = .005). Tukey’s HSD Test for multiple comparisons found that females exhibited higher blood flow in the amygdala (*p* = 0.014, 95% C.I. = -7.67, -1.42), temporal lobe (*p* < 0.005, 95% C.I. = -6.82, -1.85), anterior cingulate cortex (*p* = 0.014, 95% C.I. = -6.33, -1.17), precuneus (*p* =0.005, 95% C.I. = -9.17, -3.17), superior parietal lobe (*p* = 0.005, 95% C.I. = -6.44, -1.68), parahippocampal gyrus (*p* = 0.004, 95% C.I. = -8.27, -1.44), and hippocampus (*p* =.005, 95% C.I. = -8.11 -2.08) compared to males (Figure 2).

**Figure 2.**
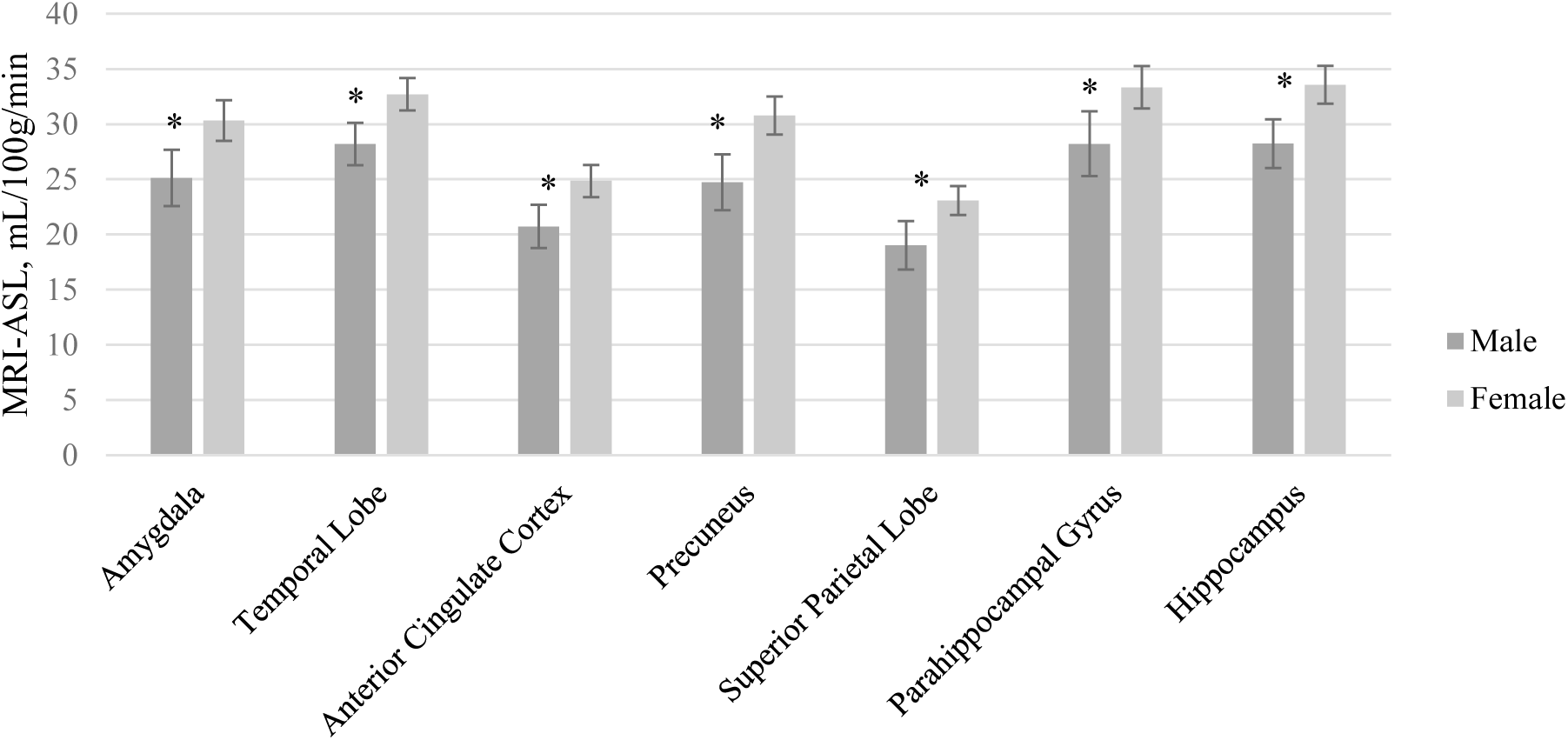
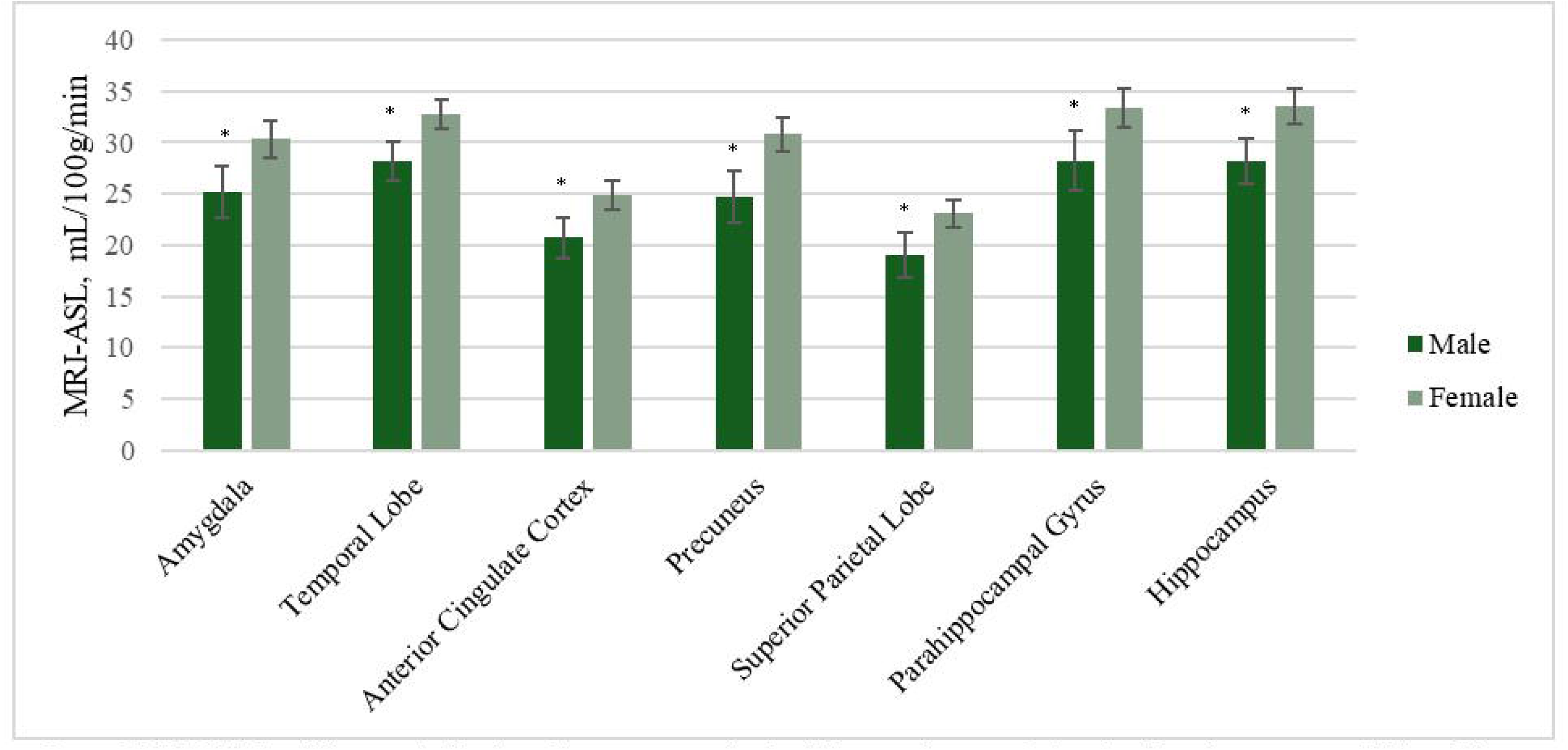
MRI-ASL Sex differences by Region of Interest accounting for differences in age and education. Error bars represent 95% confidence intervals.

### 3.3 AIM 2: ASSOCIATIONS BETWEEN SEX-DRIVEN BRAIN BIOMARKER DIFFERENCES AND AD RISK FACTORS

LASSO regressions were used to identify which risk factor variables were most strongly associated with the Aim 1 identified brain ROIs showing neuroimaging biomarker differences between sex groups. LASSO regressions selected the most informative predictors for MRI volumetric measurements of the amygdala and entorhinal cortex as well as ASL-MRI cerebral blood flow measurements of the amygdala, temporal lobe, anterior cingular cortex, precuneus, superior parietal lobe, parahippocampal gyrus, and hippocampus regions. Ten AD risk factor variables were included as potential predictors including age, education, sex, *APOE4* carrier status, non-optimal fasting plasma glucose, non-optimal mean arterial pressure, non-optimal waist-to-hip ratio, non-optimal total body fat percentage, android fat percentage, and gynoid fat percentage. To evaluate sex differences, interaction terms with each predictor multiplied by sex were included in the LASSO models (females = 1, males = 0).

#### 3.3.1 MRI VOLUMES

Overall, all LASSO models for volumetric MRI ROIs performed moderately well and did not overfit with models being able to explain a range of 63 to 77% of the variation in the values of the training data coupled with appropriate RMSE values for each model. Table 5 lists the estimated coefficients and associated R^2^ and RMSE values for each model.

**Table 5.** LASSO regression estimation coefficient results by sex and imaging modality and region of interest: MRI measures

| Modality | Group | Region ($R^2$, RMSE) | Predictor | Estimate |
| --- | --- | --- | --- | --- |
| Volumetric MRI | Female | Amygdala<br>(0.626, 0.394) | Age | -0.027 |
|  |  |  | Education | -0.036 |
|  |  |  | Fasting plasma glucose | -0.001 |
|  |  |  | Waist-to-hip ratio | -0.014 |
|  | Male |  | Age | -0.019 |
|  |  |  | Education | -0.001 |
|  | Female | Entorhinal cortex<br>(0.767, 0.471) | Age | -0.078 |
|  |  |  | Education | -0.102 |
| | | | *APOE* $\epsilon 4$ status | 0.026 |
|  |  |  | Waist-to-hip ratio | -0.010 |
|  |  |  | Mean arterial pressure | -0.103 |
|  | Male |  | Age | -0.096 |
|  |  |  | Fasting plasma glucose | -0.048 |
| | | | *APOE* $\epsilon 4$ status | -0.009 |
|  |  |  | Mean arterial pressure | 0.091 |
|  |  |  | Android fat % | -0.032 |
|  |  |  | Gynoid fat % | 0.033 |
|  |  |  | Total body fat | 0.008 |
| ASL MRI | Female | Amygdala<br>($<.001$, 6.868) | Android fat % | 1.482 |
| | | | *APOE* $\epsilon 4$ status | 0.147 |
|  | Male |  | Education | -0.164 |
|  |  |  | Fasting plasma glucose | -0.123 |
|  | Female | Temporal Lobe<br>(0.040, 6.182) | Android fat % | 1.261 |
| | Female | Anterior Cingulate Cortex<br>($<.001$, 5.549) | Android fat % | 1.322 |
| | | | *APOE* $\epsilon 4$ status | 0.007 |
| | Female | Precuneus,<br>($<.001$, 13.788) | Age | 0.397 |
|  |  |  | Education | 1.629 |
| | | | *APOE* $\epsilon 4$ status | 0.095 |
|  | Female | Superior Parietal Lobe<br>(0.030, 0.180) | Age | 0.180 |
|  |  |  | Education | 0.122 |
|  | Female | Parahippocampal Gyrus<br>(0.011, 7.334) | Android fat % | 0.704 |
|  | Female | Hippocampus<br>(0.009, 7.033) | Female sex | 1.563 |
| | | | *APOE* $\epsilon 4$ status | 0.100 |
|  | Male |  | Waist-to-hip ratio | -0.232 |

Older age predicted lower volumes in both amygdala and entorhinal cortex for both men and women. Higher education predicted lower amygdala volumes for both men and women, but only lower entorhinal volumes for women. Education was not significantly associated with entorhinal cortex volume for men. Carrying an *APOE4* allele was associated with higher entorhinal cortex volume for females, but for males, carrying a *APOE4* allele was associated with lower entorhinal cortex volume. The *APOE4* allele was not significantly associated with amygdala volumes for either males or females.

Body composition variables were associated differently with brain volumes for males and females. For females, higher waist to hip ratio was associated with lower volumes in both amygdala and entorhinal cortex. For males, body composition was not significantly associated with amygdala volume. Meanwhile, for males, lower gynoid fat and total fat were associated with lower entorhinal cortex volume, but higher android fat was associated with lower entorhinal cortex volume.

We observed sex-specific patterns for glucose levels and arterial pressure. High fasting glucose levels were associated with lower amygdala volumes for females but were not associated with amygdala volumes for males. By contrast, high fasting glucose was associated with lower entorhinal cortex volumes for males, but not for females. High mean arterial pressure was associated with lower volume in the entorhinal cortex for women and higher volume for men.

Arterial pressure was not associated with amygdala volume in males or females. The pattern of MRI volumetric association results by sex are summarized in Table 6.

**Table 6.** Pattern of MRI Volume Predictors by Sex

*Pattern of MRI Volume Predictors by Sex*
|  | Female | Male |
| --- | --- | --- |
| Lower Amygdala Volume | Older age | Older age |
|  | Higher education | Higher education |
|  | High waist/hip ratio |  |
|  | High fasting glucose |  |
| Lower Entorhinal Cortex Volume | Older age | Older age |
|  | Higher education | High fasting glucose |
| | High waist/hip ratio | *APOE* $\epsilon 4$ positive |
| | *APOE* $\epsilon 4$ negative | Low arterial pressure |
|  | High arterial pressure | Higher android fat |
|  |  | Lower gynoid fat |
|  |  | Lower total fat |

#### 3.3.2 ASL-MRI CEREBRAL BLOOD FLOW

Overall, the seven LASSO models for ASL-MRI cerebral blood flow of ROIs performed very poorly and underfit the training data by explaining a range of 0 to 1% of the variation in the values of the training data. RMSE values for each model were also unstable. Results should be interpreted with caution. Table 5 lists the estimated coefficients and associated R^2^ and RMSE values for each model.

For women only, higher android fat percentage was the variable most consistently associated with higher blood flow in the greatest number of ROIs, followed by being *APOE4* positive and older age. Higher android fat percentage was associated with greater blood flow in the amygdala, parahippocampal gyrus, temporal lobe, and anterior cingulate cortex. Being *APOE4* positive predicted greater blood flow in the amygdala, hippocampus, and precuneus for women. Older age predicted greater blood flow in the precuneus and superior parietal lobe for women.

For men, more education was associated with less blood flow in the amygdala whereas for women, more education was associated with more blood flow in the precuneus and superior parietal lobe. Non-optimal fasting glucose was associated with less blood flow in the amygdala and a higher than optimal waist to hip ratio was associated with less blood flow in the hippocampus for men only. The pattern of ASL-MRI cerebral blood flow association results by sex are summarized in Table 7.

**Table 7.** Pattern of ASL-MRI Cerebral Blood Flow Predictors by Sex

*Pattern of ASL-MRI Cerebral Blood Flow Predictors by Sex*
| **Predictors of Blood Flow** | Female | Male |
| --- | --- | --- |
| Higher Amygdala Blood Flow | Higher android fat<br>*APOE* $\epsilon 4$ positive | Lower education<br>Lower fasting glucose |
| Higher Temporal Lobe Blood Flow | Higher android fat |  |
| Higher Anterior Cingulate Cortex Blood Flow | *APOE* $\epsilon 4$ positive<br>Higher android fat | |
| Higher Precuneus Blood Flow | *APOE* $\epsilon 4$ positive<br>Older age<br>Higher education | |
| Higher Superior Parietal Lobe Blood Flow | Older age<br>Higher education |  |
| Higher Parahippocampal Gyrus Blood Flow | Higher android fat |  |
| Higher Hippocampus Blood Flow | *APOE* $\epsilon 4$ positive | Low hip/waist ratio |

## 4.0 DISCUSSION

This multimodal neuroimaging study aimed to examine sex differences in preclinical AD biomarkers among amyloid positive, cognitively unimpaired, sedentary older adults. Several key findings emerged that support and extend existing literature. First, significant sex differences in MRI T1-weighted regional volumes and ASL-MRI measured blood flow were observed in regions linked to preclinical AD pathology and the estrogen receptor network. Second, AD biomarker differences were most consistently and strongly associated with impaired fasting glucose, higher than optimal waist to hip ratio, higher android fat percentage, and *APOE4* carrier status for females, especially for MRI T1-weighted volumetric findings. Third, amyloid-β levels were comparable between females and males, likely due to the selection criteria of our study. Many of the implicated cardiometabolic risk factors are modifiable. Thus, these findings support the development of sex-specific strategies for modifiable risk factors during the AD preclinical stage.

### 4.1 PET AMYLOID-β BURDEN SEX DIFFERENCES

We did not find any associations between sex and amyloid-β burden among the anterior cingulate gyrus, lateral temporal lobe, precuneus, superior parietal lobule or global amyloid-β burden after adjusting for age, education, and *APOE4* status. It is important to note that we did not expect a large amount of variance among participants given the inclusion criteria of the primary APEX study required sub-threshold or elevated amyloid-β, which may have contributed to the lack of sex differences we observed. These results are consistent with literature using cross-sectional methodology among cognitively unimpaired older adults [98] and across the lifespan [49]. Some research suggests sex differences in amyloid-β burden may exist but occur earlier in the aging process, such as during the menopause transition [47,99]. According to the estrogen hypothesis, estrogen may reduce aggregation of amyloid-β. This suggests that the decrease of estrogen during menopause may result in greater amyloid-β burden. Future studies may benefit from exploration of the potential effect of reproductive history, including years since menopause, and its relationship with amyloid-β burden among postmenopausal women. An alternative possibility for the lack of observed sex differences in amyloid-β may relate to a lack of control for AD family history in our study, for which prior studies for have demonstrated sex differences [100]. Future studies should consider family history when examining the interaction between sex and asymptomatic cerebral β-amyloidosis.

### 4.2 MRI-VOLUMETRIC SEX DIFFERENCES

Our volumetric findings are consistent with the UK Biobank, the largest single-sample study of structural sex differences in the human brain ages 40–69 [101]. The study found males generally had larger total brain volume and among similar brain regions of interest to the present study to include the hippocampus, nucleus accumbens, amygdala, caudate nucleus, dorsal pallidum, putamen, and thalamus, after controlling for intracranial volume. Interestingly, they also found women had thicker cortices compared to men globally and among these regions, except notably the entorhinal cortex which was found to be thicker among men. Future studies should independently examine sex differences among regional cortical thicknesses, which have been shown to be phenotypically independent from structural volume measurements [102]. In a sample slightly younger than the current study, Rahman et al. 2020 and Mosconi et al. (2021) also demonstrated lower MRI gray and white matter volume among women in the amygdala compared to men. Contrary to our results, they also found women had significantly lower volumes in the hippocampus, parahippocampal gyrus, insula, and caudate in addition to the superior, middle, and orbital frontal gyrus, anterior cingulate (ACC), and putamen among 46-65 year old females compared to age-matched males [47,99]. Future studies with larger sample sizes and across age groups during post-menopause should explore the potential underlying mechanisms of sex hormones in volumetric sex differences accounting for reproductive system health history.

Other studies have found no sex differences among structural volumetric measurements of the hippocampus and amygdala among samples of healthy older adults [51,103]. Contrary to these studies, our sample included sedentary older adults with asymptomatic cerebral β-amyloidosis. Further, findings from the Buckley et al. (2018) review suggest a greater amyloid-β burden sensitivity in females relative to males such that females with asymptomatic cerebral β-amyloidosis had steeper cognitive decline and neurodegeneration compared to males [98]. Taken in context of our volumetric findings, this may suggest faster neurodegeneration among women with preclinical AD risk factors compared to men. However, the cross-sectional design of the present study limits this interpretation, and longitudinal studies are needed to further investigate this potential sex difference in AD trajectory.

### 4.3 MRI-ASL CEREBRAL BLOOD FLOW SEX DIFFERENCES

Our study found that older adult women with asymptomatic cerebral β-amyloidosis had higher blood flow in regions associated with AD pathology and estrogen receptor network ROIs compared to men. The mechanism explaining the interaction between sex and cerebral blood flow in our results remains unclear. Mosconi et al. (2021) demonstrated the role of estrogen on cerebral blood flow during the menopause stages. The study found higher cerebral blood flow in postmenopausal women compared to perimenopausal women in the supramarginal gyrus, middle and superior temporal gyrus, superior and inferior front gyrus of both hemispheres [99]. In their discussion of these findings, they proposed a compensatory reaction to glucose hypometabolism, and increased ketone metabolism after menopause as an underlying reason for the increased cerebral blood flow in these women [99]. These results could also be interpreted as suggesting that blood flow is reduced during the fluctuations of perimenopause. During post menopause, blood flow may be returning to normal or possibly higher functioning as part of the compensatory reaction. Given our postmenopausal sample was older than the Mosconi et al. 2021 study, in addition to our volumetric sex difference findings, there could be a similar compensatory process occurring for women later in the aging process.

### 4.4 ASSOCIATIONS BETWEEN SEX-DRIVEN BIOMARKER DIFFERENCES AND AD RISK *FACTORS*

Only one other study to our knowledge has evaluated associations of demographic and cardiometabolic risk factors with MRI T1 weighted volumetric sex differences in cognitively unimpaired older adults [104] and none have examined predictors of MRI-ASL cerebral flood flow. In contrast to our methodological approach, Armstrong et al. 2019 followed its sample for 20 years and measured volumetric loss using cardiovascular predictors defined differently than in our study [104]. They found that for males, hypertension and higher HDL cholesterol were protective against volume loss in the hippocampus and parahippocampal gyrus. In females, obesity (as measured by body mass index ≥30 kg/m2 vs. <30 kg/m2) was found to protect against volume loss in the temporal gray matter but hypertension was associated with steeper volumetric decline in gray matter compared to men [104]. Although we did not replicate their hypertension findings in the hippocampus or parahippocampal gyrus, we also found sex differences to include high mean arterial pressure predicted higher volume in the entorhinal cortex for males and lower volume for females.

Contrary to their study, our study measured obesity using more precise methods including waist to hip ratio, DEXA measured total fat percentage, and type of fat distribution (android and gynoid fat percentages). Our study found that for men, more android fat but less gynoid fat and total fat was associated with smaller entorhinal cortex volumes. In contrast, we found for females, waist to hip ratios greater than or equal to 0.85 was associated with smaller amygdala and entorhinal cortex volumes and more android fat predicted higher cerebral blood flow. It is worth noting that measures of body composition have shown non-linear age-related associations with cognitive performance and dementia such that body fat or higher body weight may be protective at some life stages and deleterious at other stages [105]. Thus, timing may be a critical element to understanding sex differences in the influence of cardiometabolic risk factors on the brain.

Weight distribution during aging differs between males and females and may further explain some of our findings. For males, adipose tissue slowly but consistently accrues in the trunk and abdomen during aging over time (android fat distribution). For females, prior to menopause, adipose tissues accrue in the hips and thighs (gynoid fat distribution). After menopause, gynoid fat mass stabilizes while android fat distribution steeply increases into older adulthood [106,107]. Android fat distribution is associated with visceral fat accumulation which is directly related to cardiovascular disease and metabolic disease compared to other types of fat, both of which have been recognized as risk factors for Alzheimer’s disease. Females in our sample had significantly more android fat percentage on average compared to males, suggesting postmenopausal females in our sample have more visceral fat and susceptibility for increased cardiovascular risk factors.

Our sample was sedentary by design. Research has found sedentary behavior is associated with additional risk factors for Alzheimer’s disease including impaired glucose and lipid metabolism [108]. Sedentary behavior over time may lead to abdominal obesity and insulin resistance, the most prominent underlying risk factors for cardiovascular disease and metabolic syndrome [109–113]. In addition, according to the estrogen hypothesis, postmenopausal women are also more susceptible to these cardiovascular and metabolic risk factors due to the loss of estrogen and its key protective effect for glucose transport regulation and aerobic glycolysis.

This area of aging research may provide a potential understanding of the underlying cause for the association finding between android fat percentage and waist to hip ratio with brain volume and blood flow among AD pathology and estrogen receptor network region ROIs in our sample of sedentary older adult females. More research is warranted to further explain these findings.

### 4.5 STRENGTHS AND LIMITATIONS

The current analysis was unique in that it included neuroimaging techniques that considered two of the three categories as part of the A/T/N biomarker classification scheme for AD pathology [43]. Uniquely, it adds ASL imaging, a unique imaging technique for highlighting sex and age differences in cerebral blood flow. However, the current analysis is missing consideration of neurofibrillary tangle tau biomarkers, which is commonly measured by elevated CSF phosphorylated tau and elevated NFT-tau ligand uptake on PET imaging analyses. Future studies should consider examining sex differences among all three categories of the A/T/N biomarker AD pathology scheme in preclinical AD individuals.

The current study is cross-sectional in design, and therefore causal direction of associations cannot be determined. Longitudinal studies are needed to determine if the identified sex differences are predictive of dementia or suggest healthy brain aging differences over time, especially considering changes that result from the menopause transition. The use of regression analysis to examine a potential relationship between neuroimaging biomarkers and cardiovascular risk factors in a cross-sectional study is also problematic. Neurodegeneration can only be measured using longitudinal study designs. It also cannot be determined whether blood flow is increasing or decreasing over time for either men or women. Lastly, cardiovascular risk factors and health are cumulative over time therefore analyses examining health outcomes should be controlling for health and related behaviors over a lifetime to examine potential effects in later life.

Our sample consisted of a preponderance of non-Hispanic White, educated, and high socioeconomic status older adults, diminishing the external validity of the results of the current study. The imbalance between sample size of men and women and relatively small sample size of men may have limited the sex-associated difference analyses. However, the imbalanced sample size represents the ratio of Alzheimer’s disease prevalence between men and women with two thirds of patients with AD being women. Our results are only generalizable to persons who identify as female or male and may not apply to trans-women or trans-men. Sex and gender exist on spectrums. Research on sex-differences must consider the health and aging experiences of trans-people or for persons who identify elsewhere on the sex/gender spectrum and examine factors unique to this population such as hormone therapy.

Another limitation of the current study was the binomial (yes/no) categorization of AD risk factors as predictive measures in the analysis including impaired fasting glucose, high mean arterial pressure, higher than optimal total body fat percentage, and higher than optimal waist to hip ratio. Although dichotomizing the risk factors aligned with typical approaches, we acknowledge it simplifies what is an underlying continuous process. Measurement imprecision inevitably results in some classification errors, particularly for values close to cutoff points. Future studies should consider the use of continuous biomarker variables to provide a more accurate representation of the severity of the risk factors.

The current study is limited by its lack of control for other well-established risk factors for AD with known sex and gender differences including other health conditions (e.g., dyslipidemia, hypercholesterolemia), health behaviors (e.g., diet, sleep, smoking), socioeconomic considerations (e.g., occupation, household income), and psychological factors (e.g., depression) [114–116]. Another major limitation of the current study is its lack of reproductive history consideration, specifically menopause type (e.g., natural, medically or surgically induced) and treatment (e.g., hormone therapy) as potential contributors to AD risk. Natural menopause has been associated with AD-risk-specific neuropathology including amyloid-β deposition and decreased hippocampal volume [45,46]. Further, research has identified a greater risk for AD associated with early medically or surgically induced menopause that occurs prior to natural menopause [117,118]. A recent review and meta-analysis of research on hormone replacement therapy and its relationship to cognition and AD risk indicate a lack of benefits and potential harmful effects, except in women who undergo early surgically induced menopause. Current research suggests the formulation, route of administration, and timing of hormone replacement therapy initiation produce different effects and are critical to understanding the efficacy of hormone therapy for AD prevention [15,119–130]. Future studies should consider the combination of these risk factors, especially reproductive history, among one individual and the potential sex differences among these risk factors in relation to preclinical AD biomarkers.

### 4.6 CONCLUSION

In summary, we examined associations of AD risk factors with AD biomarkers by sex in our cohort of sedentary older adults. We found widespread sex differences in T1 MRI weighted volumetrics in addition to ASL-MRI blood flow. Sedentary older adult women with asymptomatic cerebral β-amyloidosis demonstrated smaller volumes in addition to higher blood flow in AD pathology and estrogen receptor network ROIs. Genetic and chronic health risk factors were most strongly associated with these sex-related differences for the female group only including higher than optimal waist to hip ratio was most strongly associated with lower volume and higher android fat percentage and *APOE4* carrier status were most strongly associated with higher blood flow, with stronger evidence for MRI T1 weighted volumetric findings. Future investigations with larger sample sizes should include reproductive history characteristics including hysterectomy status and hormone replacement in an effort to investigate a potential underlying sex-specific biological pathway to brain aging to explain these differences, as was found among midlife women in the Rahman et al. 2020 study [47]. These findings highlight the importance of considering differences in patterns of neurodegeneration and brain blood flow among preclinical AD older adult males and females when developing effective AD interventions and prevention strategies.

## Data Availability

The data supporting the findings of this study are openly available in Harvard Dataverse at https://doi.org/10.7910/DVN/B9I1F8.

## ACKNOWLEDGEMENTS & FUNDING

This work was supported by the National Institutes of Health R01 AG043962 (JMB); K99 AG050490 (JKM) and gifts from Frank and Evangeline Thompson (JMB), The Ann and Gary Dickinson Family Charitable Foundation, John and Marny Sherman, and Brad and Libby Bergman. Institutional infrastructure support for testing was provided in part by UL1 TR000001 (RJB) and P30 AG035982 (RHS JMB). Lilly Pharmaceuticals provided a grant to support F18-AV45 doses and partial scan costs (JMB). AW is supported by an NIH grant from NIGMS and OD 1P20GM152280. MNK is supported by NIA grants T32 AG078114 and P30 AG072973.

The content is solely the responsibility of the authors and does not necessarily represent the official views of the National Institutes of Health. The funders had no role in study design, data collection and analysis, decision to publish, or preparation of the manuscript.

## CONFLICT OF INTEREST

The authors have no conflict of interest to report.

## Notes

### Competing Interest Statement

The authors have declared no competing interest.

### Author Declarations

The project was approved by the University of Kansas Medical Center Human Subjects Committee

